# Dissecting the genetic overlap between three complex phenotypes with trivariate MiXeR

**DOI:** 10.1101/2024.02.23.24303236

**Authors:** Alexey A. Shadrin, Guy Hindley, Espen Hagen, Nadine Parker, Markos Tesfaye, Piotr Jaholkowski, Zillur Rahman, Gleda Kutrolli, Vera Fominykh, Srdjan Djurovic, Olav B. Smeland, Kevin S. O’Connell, Dennis van der Meer, Oleksandr Frei, Ole A. Andreassen, Anders M. Dale

## Abstract

Comorbidities are an increasing global health challenge. Accumulating evidence suggests overlapping genetic architectures underlying comorbid complex human traits and disorders. The bivariate causal mixture model (MiXeR) can quantify the polygenic overlap between complex phenotypes beyond global genetic correlation. Still, the pattern of genetic overlap between three distinct phenotypes, which is important to better characterize multimorbidities, has previously not been possible to quantify. Here, we present and validate the trivariate MiXeR tool, which disentangles the pattern of genetic overlap between three phenotypes using summary statistics from genome-wide association studies (GWAS). Our simulations show that the trivariate MiXeR can reliably reconstruct different patterns of genetic overlap. We further demonstrate how the tool can be used to estimate the proportions of genetic overlap between three phenotypes using real GWAS data, providing examples of complex patterns of genetic overlap between diverse human traits and diseases that could not be deduced from bivariate analyses. This contributes to a better understanding of the etiology of complex phenotypes and the nature of their relationship, which may aid in dissecting comorbidity patterns and their biological underpinnings.

**Availability and implementation:** The trivariate MiXeR tool and auxiliary scripts, including source code, documentation and examples of use are available at https://github.com/precimed/mix3r

## Introduction

Many human traits and disorders are highly polygenic, with thousands of associated loci discovered to date [1]. Most functional genetic loci affect multiple phenotypes spawning intricate patterns of genetic overlap among them [2]. Characterization of patterns of overlapping genetic architecture across multiple phenotypes has generated key discoveries in human genetics in recent years. This characterization has helped reveal the shared genetic underpinnings of a wide range of common human diseases [3]. Ultimately, a more complete understanding of the complex genetic relationships between various traits and disorders can help to elucidate mechanisms underlying multimorbidity [4], which is an increasing global health challenge [5]. It can also lead to improvements in disease classification and diagnostics.

Previous causal mixture model (MiXeR) tools have been implemented to model genetic architecture and characterize genetic overlap. Univariate MiXeR [1] was developed to quantify characteristics of genetic architecture for complex phenotypes, including polygenicity, which reflects the number of genetic variants influencing a phenotype. This approach was extended to bivariate MiXeR [6] to quantify the overlapping polygenic components between two phenotypes regardless of the effect directions. While genetic correlation is often used to assess genetic overlap between complex phenotypes, its ability to detect overlap is limited to pairs of phenotypes where the bulk of variants have either concordant or discordant directions of effects. A pair of phenotypes sharing variants with a balanced mixture of concordant and discordant effects will provide a genetic correlation close to zero, similar to a pair of genetically disjoint phenotypes, making these two scenarios indistinguishable [7]. Bivariate MiXeR has helped to better characterize the relationship between various pairs of phenotypes beyond genetic correlation, effectively capturing shared genetics with mixed directions of effects [3, 8-10]. However, neither genetic correlation nor bivariate MiXeR can directly estimate genetic overlap across three phenotypes. Apart from trivial cases of non-overlapping or completely overlappling phenotypes, trivariate overlap cannot be reconstructed from a series of three bivariate analyses (analyzing each pair of phenotypes within a triad). Indirect reconstruction of trivariate overlap from three bivariate analyses under a naïve assumption of a maximum entropy probability distribution of overlapping parts may lead to erroneous estimates. For a given combination of three bivariate overlaps, with no additional prior knowledge, the maximum entropy distribution of the trivariate overlap can be conceptualized as the center of all possible trivariate overlap distributions. Investigating genetic overlap among triads of phenotypes can help reveal important aspects of genetic overlap associated with different scenarios of overlap between phenotypes, tissue types or biological mechanisms.

Here, we present trivariate MiXeR, which disentangles the pattern of polygenic overlap among three complex phenotypes using summary statistics from genome-wide association studies (GWAS). We first conduct a series of simulations covering diverse scenarios of genetic overlap among three phenotypes and demonstrate that the tool can reliably reconstruct different patterns of trivariate genetic overlap. We then apply trivariate MiXeR to GWAS summary statistics for eight complex phenotypes representing a range of human traits and disorders for which epidemiological studies suggest shared causal pathways. Our analyses demonstrate non-trivial patterns of trivariate genetic overlap that are substantially different from naïvely expected patterns derived from three bivariate analyses following the principle of maximum entropy.

## Methods

### Trivariate MiXeR model

The method extends the bivariate MiXeR model [6] for the case of three phenotypes, leaving the basic assumptions of the model unchanged. Briefly, an additive model of genetic effects is considered. In the univariate analysis, the direct (not induced by linkage disequilibrium) effect *β*_*j*_ of the *j*^th^ variant on a phenotype is modeled as a mixture of null and phenotype-influencing components characterized by two parameters: the proportion of variants influencing the phenotype (polygenicity, *π* ∈ [0,1]) and the variance of their effect sizes (discoverability, *σ*^2^):

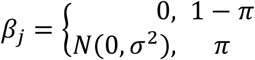

where *N*(0, *σ*^2^) is a normal distribution with zero mean and *σ*^2^ variance.

In a joint analysis of three phenotypes (*i* = 1, 2, 3), a fraction of variants may affect all three phenotypes (*π*_123_), other variants may affect a pair of phenotypes but not the third phenotype (*π*_12_, *π*_13_, *π*_23_), some variants might be phenotype-specific (*π*_1_, *π*_2_, *π*_3_) while most variants are expected to have no effect on any phenotype (*π*_0_ = 1 − *π*_1_ − *π*_2_ − *π*_3_ − *π*_12_ − *π*_13_ − *π*_23_ − *π*_123_). We assume all variants for a given phenotype follow the same distribution of effect sizes (with corresponding discoverabilities 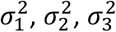), regardless of their effects on the other two phenotypes. Genetic correlations are modeled by introducing correlations of effect sizes within each of three pairwise overlaps (*ρ*_12_, *ρ*_13_, *ρ*_23_). With these assumptions, the trivariate distribution of direct effects of the j^th^ variant is modeled as a mixture of eight components:

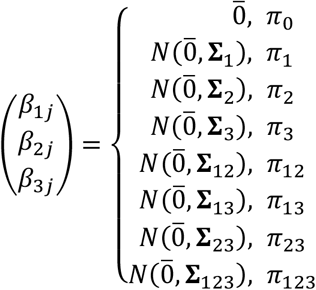

where 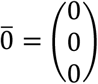 and 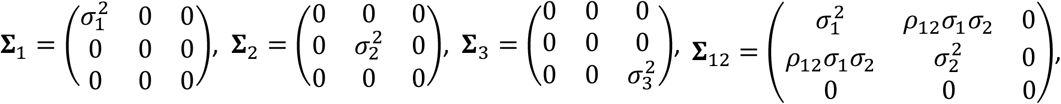 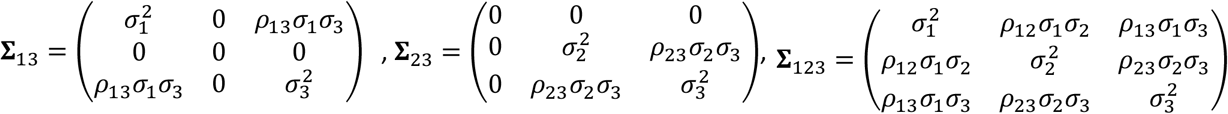 are covariance matrices of multivariate normal distributions corresponding to the different phenotype-influencing components.

The joint signed association test statistics (z-score) of the j^th^ variant is then given by:

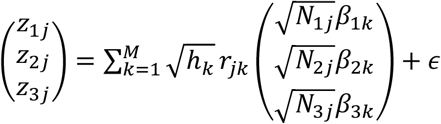

where *N*_*ij*_ (*i* = 1, 2, 3) is the sample size of the GWAS for the i^th^ phenotype and j^th^ variant, *h*_*k*_ is the heterozygosity of variant *k, M* is the number of variants in linkage disequilibrium (LD) with the variant *k,r*_*jk*_ is the Pearson’s correlation coefficient between the genotypes of the j^th^ and k^th^ variants (quantifying LD), and 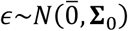 is a normally distributed vector of residuals with covariance matrix

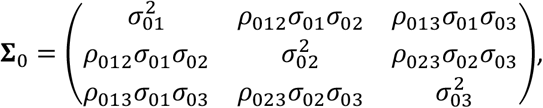

where 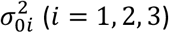 is a residual variance of the i^th^ phenotype and *ρ*_0*i*_ (*i, j* = 1, 2, 3) is a correlation between residuals of the i^th^ and j^th^ phenotypes. Nineteen parameters of the model (*π*_1_, *π*_2_, *π*_3_, *π*_12_, *π*_13_, *π*_23_, *π*_123_, *σ*_1_, *σ*_2_, *σ*_3_, *σ*_01_, *σ*_02_, *σ*_03_, *ρ*_12_, *ρ*_13_, *ρ*_23_, *ρ*_012_, *ρ*_013_, *ρ*_023_) are estimated by maximizing the likelihood of the z-scores observed in the GWAS summary statistics using a step-wise procedure. First, three univariate analyses are performed to estimate univariate polygenicities 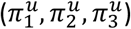, discoverabilities (*σ*_1_, *σ*_2_, *σ*_3_) and residual variances (*σ*_01_, *σ*_02_, *σ*_03_) for each of the three phenotypes. Then the bivariate analyses are performed to estimate pairwise genetic overlaps 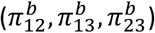, correlations of effect sizes within each of the three pairwise overlaps (*ρ*_12_, *ρ*_13_, *ρ*_23_) and correlations between residuals (*ρ*_012_, *ρ*_013_, *ρ*_023_) for each of the three pairs of phenotypes with univariate parameters fixed to the values obtained at the univariate step. Finally, the genetic overlap between all three phenotypes (*π*_123_) is estimated with both univariate and bivariate parameters fixed to the values obtained in the univariate and bivariate steps. Phenotype pair-specific polygenicities can then be calculated as:

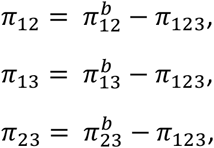

and phenotype-specific polygenicities can be calculated as:

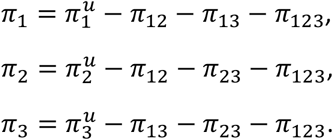

Nomenclature of pattern proportion parameters (*π*_1_, *π*_2_, *π*_3_, *π*_12_, *π*_13_, *π*_23_, *π*_123_) is illustrated in Figure 1 (row 3, column B).

**Figure 1.**
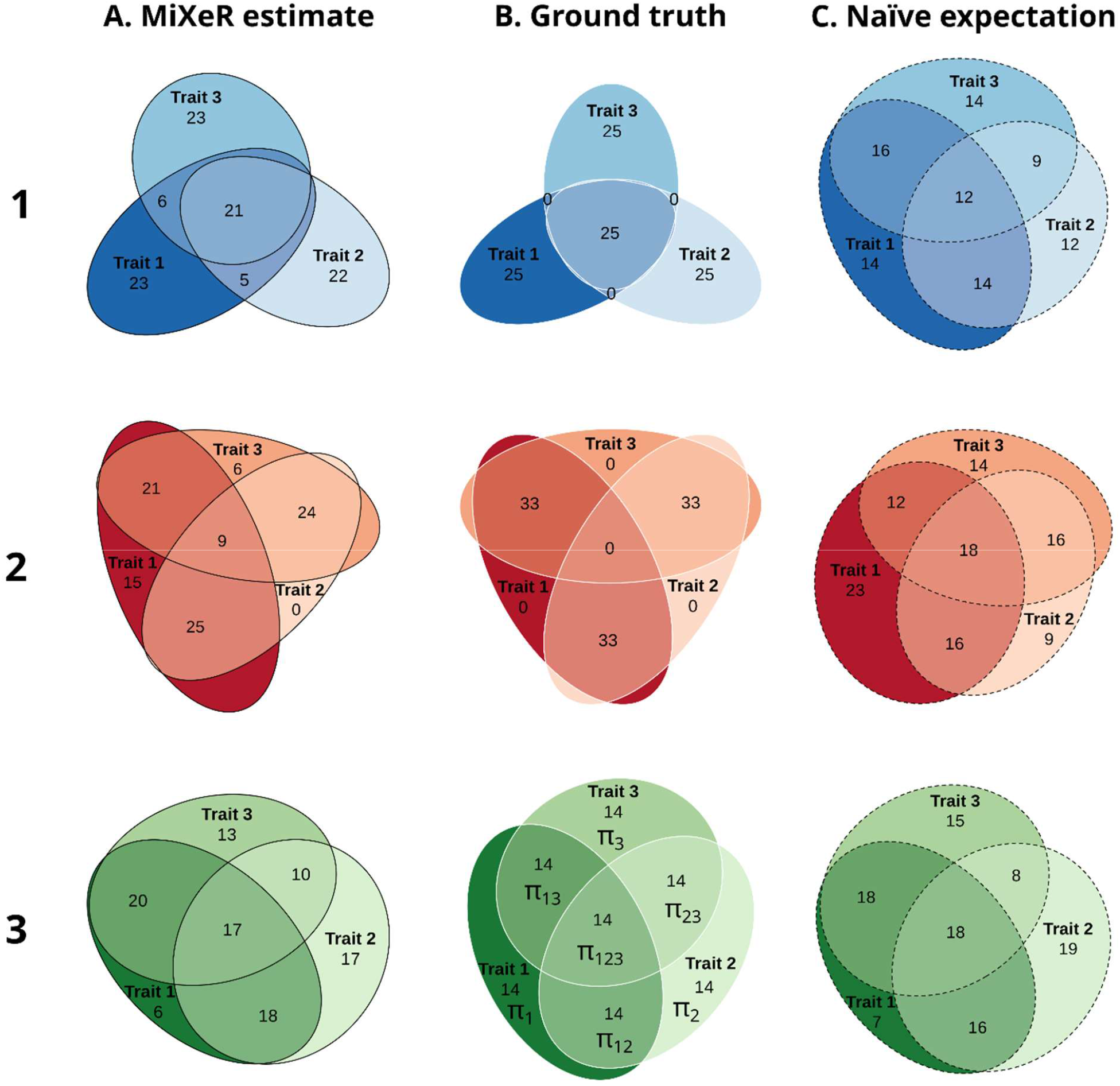
Simulated data. Three different scenarios of genetic overlap in simulated data (rows) estimated by trivariate MiXeR (column A, solid black outline), compared to the theoretical true pattern of the simulated overlap (column B, solid white outline) and the overlap pattern expected for the estimated bivariate overlaps under naïve assumption of maximum entropy (column C, dashed black outline). Row 1 (blue colors): “Core” scenario; Row 2 (red colors): “Ring” scenario; Row 3 (green colors): “Equilibrium” scenario (see ‘Methods’ for further details). For each simulation scenario (within each row), for every area of each diagram, its percentage with respect to the combined total area of all three phenotypes in the estimated diagram (Column A) is shown (rounded to the closest integer), i.e., percentages within each diagram in the column A add up to 100 and percentages within each row are directly comparable with those shown in column A. Since percentages in column C are also given with respect to the combined total area of the corresponding diagram in column A, the sum of percentages in column C is not necessarily equal to 100. For each scenario phenotypes were simulated independently, therefore “Trait 1”, “Trait 2” and “Trait 3” in row 1 are not the same as “Trait 1”, “Trait 2” and “Trait 3” in row 2 or in row 3 of the figure respectively. The middle diagram in the bottom row (row 3, column B) shows the nomenclature for proportions of areas within the pattern (*π*_1_, *π*_2_, *π*_3_, *π*_12_, *π*_13_, *π*_23_, *π*_123_) used throughout the text.

Univariate, bivariate and trivariate log-likelihood functions are implemented using numerical integration of the characteristic function applying a trapezoidal rule with fixed step size as described previously [11].

### Simulation setup

To validate the method and to test its ability to discriminate different scenarios of genetic overlap, we performed a series of analyses with simulated data. GWAS summary statistics for simulations were generated based on participants randomly selected from the UK Biobank using 100,000 unrelated (defined by 22020 data-field) white British (defined by 22006 data-field) individuals and version 3 of the genetic data. UK Biobank data was obtained under accession number 27412. Autosomal variants with minor allele frequency above 0.1%, genotype missingness below 10%, imputation info score above 0.8 and passing Hardy-Weinberg equilibrium test at p=1E-10, totaling 12,926,691 variants were included in the analysis. A set of quantitative phenotypes with equal polygenicity (*π* = 0.002), equal SNP-heritability (*h*2 = 0.4) and different patterns of genetic overlap were generated using the SIMU tool [12]. For each phenotype, a given number of phenotype-influencing variants (*n* = 25,853 ≈ 12,926,691 ∗ 0.002) were selected at random. The effect sizes for the selected variants were sampled from a standard normal distribution and scaled to obtain the predefined SNP-heritability. For each individual analyzed, a quantitative synthetic phenotype was then generated as the sum of allelic effects over all phenotype-influencing variants complemented by a certain proportion of random Gaussian noise (representing environmental effects) required to keep the predefined level of heritability. Association analysis was performed using PLINK2 [13, 14] with sex, age and the first 10 genetic principal components included as covariates. Three simulation scenarios were considered:

1. “Core”: only triple overlap, i.e., all overlapping variants are shared between all three phenotypes, for each phenotype half of phenotype-influencing variants also influence both other phenotypes. *π*_1_ = *π*_2_ = *π*_3_ = *π*_123_, *π*_12_ = *π*_13_ = *π*_23_ = 0. Presented in Figure 1, row 1.
2. “Ring”: no triple overlap, for each phenotype half of phenotype-influencing variants also influence one of the remaining two phenotypes and the second half influences another phenotype, *π*_12_ = *π*_13_ = *π*_23_, *π*_1_ = *π*_2_ = *π*_3_ = *π*_123_ = 0. Presented in Figure 1, row 2.
3. “Equilibrium”: balanced mixture of all three phenotypes, *π*_1_ = *π*_2_ = *π*_3_ = *π*_12_ = *π*_13_ = *π*_23_ = *π*_123_. Presented in Figure 1, row 3.

For each triad of phenotypes, sixteen independent optimization runs were performed to maximize the likelihood of the GWAS z-scores observed in different subsets of 500,000 randomly selected variants. We then calculated the median across these sixteen runs for each polygenicity parameter 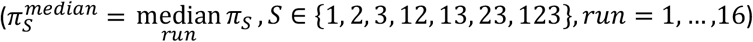 and find the run with the smallest deviation from the median overlap pattern. A Euler diagram for this run is then presented both for the simulated data and for the real data analysis. Of note, the pattern constituted from the median polygenicities 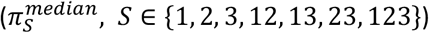 is not guaranteed to be feasible itself, since proportions in the overlap of three phenotypes are constrained, as described in the “Naïve expectation” section below, and these constraints are not necessarily fulfilled for the median proportions across multiple patterns which are themselves feasible.

### Genome-wide association studies (GWAS) data

For the analysis of the real data, we used publicly available GWAS summary statistics on eight phenotypes: ulcerative colitis [15], psoriasis [16] (FinnGen, release 9, phenotypic code L12_PSORIASIS), multiple sclerosis [17], type 2 diabetes [18], estimated glomerular filtration rate [19], high-density lipoprotein [20], placental weight (fetal GWAS adjusted for fetal sex and gestational duration) [21], height [22] and schizophrenia [23]. All summary statistics were based on individuals of European ancestry. We analyzed patterns of genetic overlap for three triads: (1) type 2 diabetes, estimated glomerular filtration rate and high-density lipoprotein (2) ulcerative colitis, multiple sclerosis and psoriasis, (3) placental weight, height and schizophrenia. The selection of these triads is guided by epidemiological and clinical evidence and intends to illustrate that the method may provide insights into a wide spectrum of traits and disorders.

### Naïve expectation

Given univariate estimates of polygenicity for three phenotypes 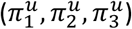, three bivariate estimates of genetic overlap between these phenotypes 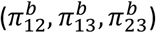 obtained in univariate and bivariate analyses for the triad of phenotypes the genetic overlap between these three phenotypes is constrained by bounds 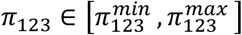, where 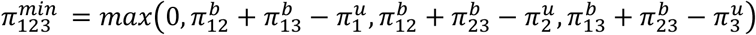 and 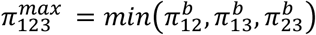. Without any prior knowledge about genetic relationships between analyzed phenotypes, a naïve expectation about the value of *π*_123_ follows the principle of maximum entropy:

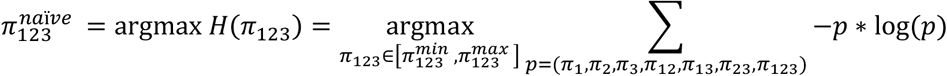

In other words for the given univariate 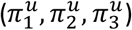 and bivariate 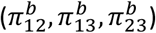 polygenicities, the naïve trivariate polygenicity 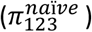 is selected so that among all the possible distributions (*π*_1_, *π*_2_, *π*_3_, *π*_12_, *π*_13_, *π*_23_, *π*_123_) with the constraint’s 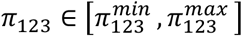, the probability distribution 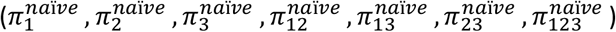 has maximum entropy, where

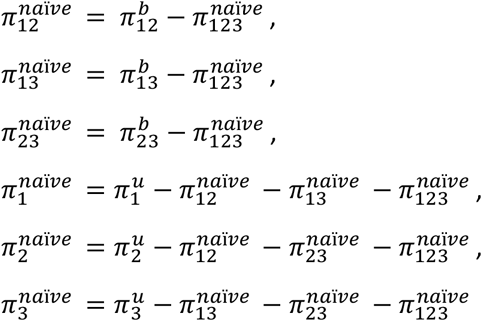

are the naïvely expected phenotype pair-specific and phenotype-specific polygenicities. Of note, polygenicities for the naïve expectation 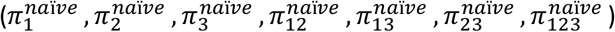 are calculated based on univariate 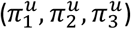 and bivariate 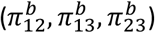 polygenicities estimated by the trivariate MiXeR model, therefore the univariate polygenicity of each phenotype as well as the shared polygenicity of each phenotypic pair are the same for the naïvely expected pattern and the pattern estimated by the trivariate MiXeR, i.e.,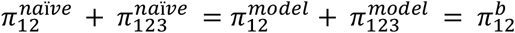, and similar for other bivariate and univariate polygenicities.

### Linkage disequilibrium reference panel

Both simulations and real data analyses were performed using an LD reference panel constructed based on 10,000 randomly selected unrelated white British individuals from the UK Biobank. Quality control procedures for variants were identical to those described in the ‘Simulations’ section leaving 12,926,691 variants in the LD reference panel. PLINK 1.9 [13, 24] was applied to estimate r2 coefficients within each autosome using --ld-window 1000000 --ld-window-kb 20000 --ld-window-r2 0.01 parameters. The resulting text files were then processed to produce input files in the format required by MiXeR, using the scripts provided in the code repository.

### Trivariate MiXeR model implementation

Building on the same assumptions as the bivariate MiXeR model, we have extended the framework to include three phenotypes modeling the trivariate distribution of genetic effects as a mixture of eight components. Compared to the bivariate MiXeR tool, the code for log-likelihood estimation was re-implemented using numerical integration of the characteristic function. This facilitated more stable convergence of the optimization algorithm and reduced fluctuations in the estimates caused by the random sampling approach applied in the bivariate MiXeR v1.3 implementation [25], at the cost of substantially increased computational burden. To cope with the increased computational demand, performance-critical parts have been accelerated using GPUs. Trivariate MiXeR code can only be deployed on machines with GPUs supporting NVIDIA CUDA, which are now commonly available on high-performance computing (HPC) facilities or cloud computing facilities.

The trivariate MiXeR tool is implemented in Python mainly using numpy [26], scipy [27] and pandas [28] packages, while the ‘numba’ just-in-time (JIT) compiler [29] is used to translate the Python and numpy-based routines into machine code. Performance-critical steps rely on the availability of a graphics processing unit (GPU, NVIDIA CUDA). Overlapping patterns are visualized using the ‘eulerr’ R package [30]. The execution environment with all dependencies can be created using Conda [31] mamba or micromamba [32].

Input parameters for the analysis can be tuned and provided in the configuration file in JSON format. An example configuration file showing parameters is available in the code repository (https://github.com/precimed/mix3r/blob/main/config_t2d_hdl_egfr_oct30_1.paper.json). Important parameters used in all presented analyses are: maf_thresh = 0.05 – z-scores of the variants with minor allele frequency (MAF) below 5% were not used for optimization (MAF is estimated from the same genotypes which are used to construct the LD reference panel); info_thresh = 0.8 – z-scores of the variants with imputation INFO score below 0.8 are not used for optimization, if input sumstats do not contain INFO column the filter is ignored; z_thresh = 32 – z-scores with absolute value larger than 32 were not used for optimization; exclude_regions = 6:25000000-34000000 – variants from the major histocompatibility complex (chr6:25000000-34000000, hg19 genomic guild) were not used for optimization; do_pruning = true, r2_prune_thresh = 0.8 – prior to optimization, variants were randomly pruned with allelic correlation threshold r2 < 0.8; n_random = 500000 – a subset of 500K variants was randomly selected for optimization from all variants surviving random pruning; rand_prune_seed = 1 – a seed for the generator of the pseudo-random numbers (controls both random pruning and random sub-setting of variants), changing this parameter while keeping all other parameters unchanged allows the user to repeat the analysis with a different subset of variants. In this study rand_prune_seed = 1, …, 16 were used to produce 16 independent runs for each triad of phenotypes.

## Results

### Application to simulated data

Our simulations demonstrated that MiXeR was able capture the true patterns of genetic overlap for various simulated scenarios (Figure 1). For “core” (Figure 1, row 1) and “ring” (Figure 1, row 2) scenarios, the trivariate MiXeR model accurately captured the disproportional pattern of overlap in the simulated datasets (column A), while the result expected for the estimated bivariate overlaps under the naïve assumption of the maximum entropy probability distribution (column C) revealed substantial deviation from the true simulated pattern (column B). For the balanced “equilibrium” scenario (Figure 1, row 3) the true pattern (column B) followed the maximum entropy principle thus the naïve expectation (column C) should be no worse than the pattern reconstructed by the trivariate MiXeR (column A). As can be seen in this case the naïve pattern and the pattern reconstructed by the trivariate MiXeR model were very similar, illustrating adequate model fit. Estimates of all model parameters for 16 independent MiXeR runs for “core”, “ring” and “equilibrium” scenarios are shown in Tables S1, S2 and S3 respectively.

### Application to real data

We applied trivariate MiXeR to GWAS summary statistics on three triads of phenotypes and compared the pattern of genetic overlap estimated by trivariate MiXeR (Figure 2, column A) with the pattern expected for the given bivariate overlaps under a naïve expectation of maximum entropy probability distribution (Figure 2, column B).

**Figure 2.**
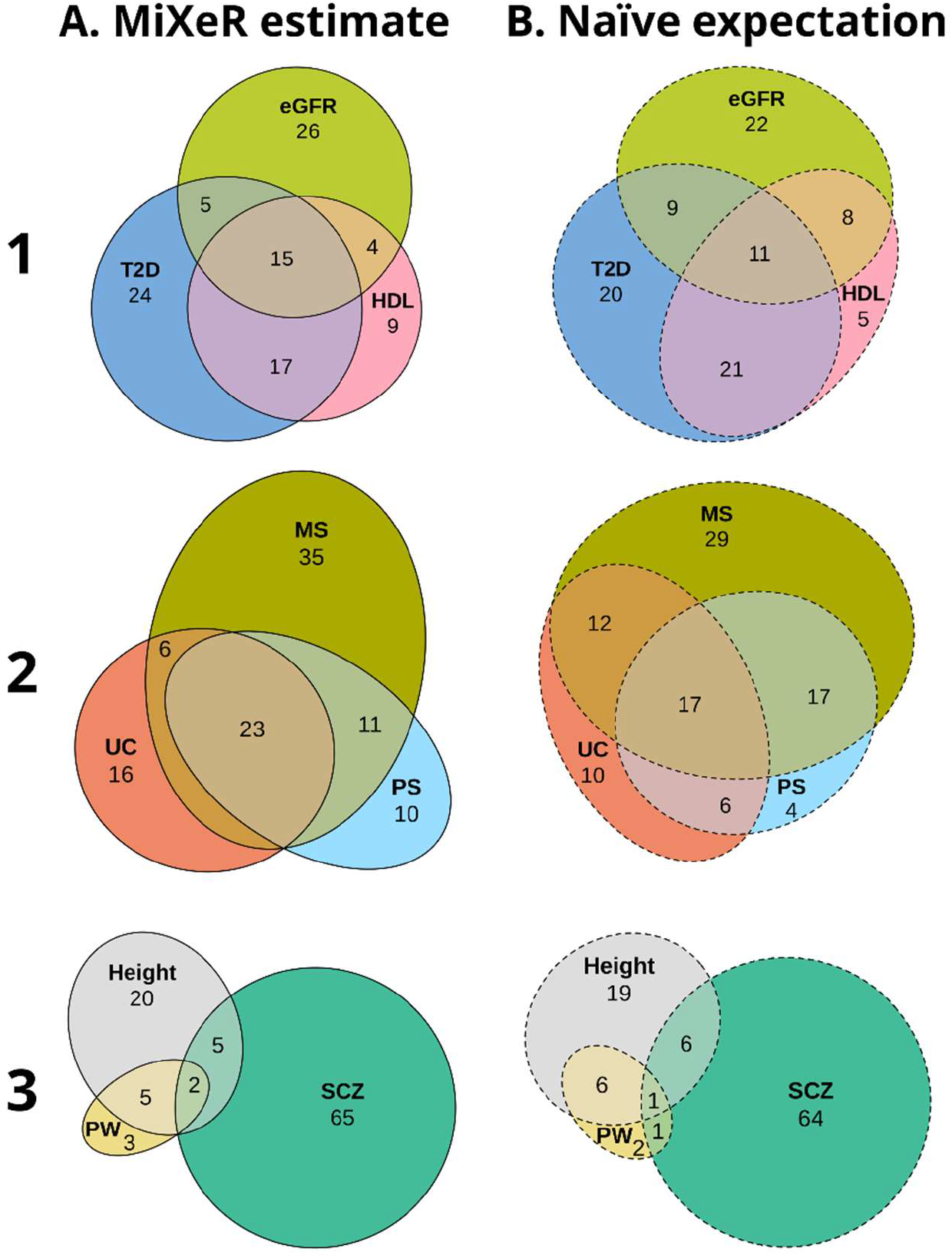
Real data. Genetic overlap between selected human traits and disorders estimated by trivariate MiXeR (column A) compared to naïve expectation following the principle of maximum entropy (column B). The pattern of genetic overlap between type 2 diabetes (T2D), estimated glomerular filtration rate (eGFR) and high-density lipoprotein (HDL) (Row 1), ulcerative colitis (UC), psoriasis (PS), and multiple sclerosis (MS) (Row 2), and placental weight (PW), height, and schizophrenia (SCZ) (Row 3) demonstrate the range of differences between trivariate MiXeR estimates and naïve expectation from bivariate MiXeR. For each triad of phenotypes (within each row), for every area of each diagram, its percentage with respect to the combined total area of all three phenotypes in the estimated diagram (column A) is shown (rounded to the closest integer), i.e., percentages within each diagram in column A add up to 100 and percentages within each row are directly comparable. Since percentages in column B are also given with respect to the combined total area of the corresponding diagram in column A, the sum of percentages in column B is not necessarily equal to 100.

The first analysis (Figure 2, row 1) included phenotypes with similar polygenicities: type 2 diabetes (univariate polygenicity, *π*^*u*^ = 5.2E-4), estimated glomerular filtration rate (*π*^*u*^ = 4.3E-4) and high-density lipoprotein (*π*^*u*^ = 3.8E-4). The estimated overlap between all three phenotypes substantially exceed the expected overlap constituting 15% of the combined total area (the naïve expectation 11%). Similarly, estimated phenotype-specific areas were larger than expected, comprising 24% (the naïve expectation 20%), 26% (22%) and 9% (5%) of the combined total area for type 2 diabetes, estimated glomerular filtration rate and high-density lipoprotein, respectively. The increase of triple overlap and phenotype-specific areas in MiXeR-derived patterns is compensated by the reduction of the phenotype pair-specific areas constituting 5% (9%) for type 2 diabetes and estimated glomerular filtration rate, 4% (8%) for estimated glomerular filtration rate and high-density lipoprotein, and 17% (21%) for type 2 diabetes and high-density lipoprotein pairs respectively. All model parameters are presented in Table S4.

In the second use case (Figure 2, row 2), the trivariate MiXeR analysis of ulcerative colitis (*π*^*u*^ = 1.2E-4), multiple sclerosis (*π*^*u*^ = 2.0E-4) and psoriasis (*π*^*u*^ = 1.2E-4) compared to the naïve expectation showed larger components shared between all three phenotypes (23 *vs*. 17%) and phenotype-specific components (ulcerative colitis: 16 vs 10%; multiple sclerosis: 35 *vs*. 29%, psoriasis: 10 *vs*. 4%), while phenotype pair-specific fractions were smaller (ulcerative colitis and multiple sclerosis: 6 *vs*. 12%, ulcerative colitis and psoriasis: 0 *vs*. 6%; multiple sclerosis and psoriasis: 11 *vs*. 17%). All model parameters are presented in Table S5.

The third analysis (Figure 2, row 3) included three phenotypes with different polygenicities: schizophrenia (*π*^*u*^ = 2.9E-3, constituting 72% of the combined total area), height (*π*^*u*^ = 1.3E-3, 32% of the combined total area) and placental weight (*π*^*u*^ = 3.9E-4, 10% of the combined total area), where the pattern derived from trivariate MiXeR (Figure 2, row 3, column A) is similar to the naïve expectation (Figure 2, row 3, column B). All model parameters are presented in Table S6.

The three examples illustrate the range of observed discrepancies between naïve expectations based on bivariate MiXeR and patterns estimated by trivariate MiXeR, which demonstrate the importance of applying trivariate analysis when studying the genetic architecture of multimorbidities.

## Discussion

We have developed and validated the trivariate MiXeR tool to disentangle the pattern of genetic overlap among three complex phenotypes using genome-wide data. The simulations showed that the tool can reliably reconstruct different patterns of genetic overlap. Furthermore, we have demonstrated how trivariate MiXeR can estimate the proportions of polygenic overlap among diverse human traits and diseases, highlighting patterns of genetic overlap that could not be deduced from bivariate MiXeR.

Pairwise genetic overlaps between multiple phenotypes have been extensively studied and have provided valuable insights into the shared and phenotype-specific genetic architectures of different traits and disorders [3, 33, 34]. However, estimating pairwise genetic overlaps among three phenotypes does not provide a complete picture of the genetic overlap among those three phenotypes. We show that the trivariate MiXeR model can dissect the pattern of genetic overlap among three complex phenotypes, using GWAS summary statistics, and reveal patterns of overlap that are distinct from naïve expectations based on bivariate MiXeR. We provide three examples with real phenotypes demonstrating that trivariate MiXeR can elucidate situations where a triad of phenotypes can overlap disproportionately, providing novel insights into the variability in overlapping genetic underpinnings among those phenotypes.

In clinical and epidemiological studies, type 2 diabetes has been associated with structural changes and abnormal function of high-density lipoprotein [35], which may impact renal function and increase the risk of kidney disease [36, 37]. Our trivariate MiXeR analysis of type 2 diabetes, high-density lipoprotein and a renal function measure (estimated glomerular filtration rate) demonstrates substantial polygenic overlap between these three phenotypes that were different from the overlap pattern expected from bivariate MiXeR results. These results nevertheless show a mixture of trivariate and bivariate overlapping polygenic components characterizing the shared genetic architecture of these phenotypes and suggesting a complex genetic relationship.

Our analysis of genetic overlap between ulcerative colitis, psoriasis and multiple sclerosis revealed a large number of disease-specific variants while the shared component was predominantly within the triple overlap. These findings are consistent with the hypothesis that there is a common genetic basis for immune-linked diseases [38] with a core combination of genetic mutations [39]. Disturbance of these hypothetical core immune processes might activate the breakdown in immune tolerance [38] necessary to trigger any of these diseases. The subsequent developmental trajectory of a given autoimmune disease may then be driven, in part, by disease-specific genetic factors. Determining and disentangling core and specific genetic factors for immune diseases might provide valuable insights into key immune pathways and cell types involved in disease mechanisms, with the potential for drug target development.

There is a growing interest in the role of the placenta in neurodevelopment and the onset of later psychiatric disorders [40, 41] but evidence for a genetic link remains elusive [21]. Our analysis of genetic overlap between placental weight, adult height and schizophrenia shows that placental weight shares a considerable fraction of its genetic underpinnings with height, while its genetic overlap with schizophrenia is modest and is also common with height. Sporadic genetic overlap is expected for polygenic phenotypes and might represent overlapping core regulatory and house-keeping genes involved in critical processes within multiple cell types. The observed pattern of genetic overlap may therefore indicate a predominance of non-specific genetic overlap between placental weight and schizophrenia.

The development of advanced methods to obtain deeper insight into the genetic overlap among multiple traits and disorders may contribute to improvements in disease nosology. Applying trivariate MiXeR to existing diagnostic categories, sub-phenotypic or symptom-level measures may inform the revision of current classification systems and provide novel insights into the nosological relationship between complex human disorders [42, 43]. Further, the triangulation of overlapping genetic patterns between disorders, biological markers and instrumental variables may inform theories regarding the potential biological mechanisms underlying multimorbidity patterns.

Trivariate MiXeR has the same limitations as the univariate and bivariate MiXeR models [1, 6]. The underlying model is sensitive to LD structure estimation and the reliability of parameter estimates depends on the statistical power of the input GWAS summary statistics. The model makes several simplifying assumptions, including uniform distribution of phenotype-influencing variants across the genome and the effect size’s independence from allele frequency, LD, and location in the genome. These assumptions may be violated to different degrees for different phenotypes, making the model less suitable for some phenotypes than for others. Analysis of phenotypes combining a handful of extremely strong genetic effects with a weak polygenic background (for example Alzheimer’s disease [1]) may be sensitive to the selection of variants used for parameter fitting. To assess the stability of optimization convergence and robustness of obtained results we perform multiple independent runs using different subsets of variants and carefully assess the variation of parameter estimates across runs to evaluate the model’s suitability for each set of phenotypes.

In conclusion, we have developed and validated the trivariate MiXeR model and demonstrated its utility in disentangling the pattern of genetic overlap between three phenotypes using GWAS summary statistics. We provide the tool implementing the model along with documentation and examples of how to use it. The trivariate MiXeR can help to provide a better understanding of the genetic relationship between complex polygenic phenotypes with particular relevance to multimorbidities.

## Data Availability

All data produced in the present work are contained in the manuscript.

https://github.com/precimed/mix3r

## Acknowledgements

This research has been conducted using data from the UK Biobank, a major biomedical database (http://www.ukbiobank.ac.uk). We want to acknowledge the participants and investigators of the FinnGen study. This work was performed on the TSD (Tjeneste for Sensitive Data) facilities, owned by the University of Oslo, operated, and developed by the TSD service group at the University of Oslo, IT-Department (USIT). Computations were also performed on resources provided by UNINETT Sigma2—the National Infrastructure for High Performance Computing and Data Storage in Norway.

