## Supplementary tables for "Dissecting the genetic overlap between three complex phenotypes with trivariate MiXeR"

**Table S1.** Parameter estimates for 16 independent MiXeR runs for the "core" simulated scenario.
The first column show the index of the optimization run (1-16), with the run having the smallest deviation from the median overlap pattern (as described in the methods section) marked with asterisk. Other columns show univariate polygenicities ($\pi_1^u, \pi_2^u, \pi_3^u$), discoverabilities ($\sigma_1, \sigma_2, \sigma_3$) and residual variances ($\sigma_{01}, \sigma_{02}, \sigma_{03}$) for Trait 1, Trait 2 and Trait 3 respectively; pairwise (bivariate) genetic overlaps ($\pi_{12}^b, \pi_{13}^b, \pi_{23}^b$), correlations of effect sizes within each of the three pairwise overlaps ($\rho_{12}, \rho_{13}, \rho_{23}$) and correlations between residuals ($\rho_{012}, \rho_{013}, \rho_{023}$) for Trait 1 and Trait 2, Trait 1 and Trait 3 and Trait 2 and Trait 3 pairs respectively; the genetic overlap between all three phenotypes ($\pi_{123}$).

| Index | $\pi_1^u$ | $\sigma_2$ | $\sigma_{01}$ | $\pi_2^u$ | $\sigma_2$ | $\sigma_{02}$ | $\pi_3^u$ | $\sigma_3$ | $\sigma_{03}$ | $\pi_{12}^b$ | $\rho_{12}$ | $\rho_{012}$ | $\pi_{13}^b$ | $\rho_{13}$ | $\rho_{013}$ | $\pi_{23}^b$ | $\rho_{23}$ | $\rho_{023}$ | $\pi_{123}$ |
|---|---|---|---|---|---|---|---|---|---|---|---|---|---|---|---|---|---|---|---|
| 1 | 2.23E-03 | 8.40E-05 | 1.039 | 1.81E-03 | 1.04E-04 | 1.029 | 2.07E-03 | 9.01E-05 | 1.026 | 1.03E-03 | 0.000 | 0.000 | 1.08E-03 | 0.009 | -0.012 | 8.86E-04 | 0.053 | 0.003 | 8.86E-04 |
| 2 | 2.20E-03 | 8.63E-05 | 1.034 | 2.01E-03 | 1.00E-04 | 1.017 | 1.91E-03 | 9.81E-05 | 1.031 | 1.07E-03 | 0.000 | 0.000 | 1.03E-03 | 0.003 | -0.008 | 8.28E-04 | 0.068 | 0.002 | 8.28E-04 |
| 3 | 2.30E-03 | 8.56E-05 | 1.025 | 1.85E-03 | 1.03E-04 | 1.028 | 1.92E-03 | 9.96E-05 | 1.028 | 1.14E-03 | 0.000 | 0.001 | 1.02E-03 | 0.005 | -0.013 | 7.18E-04 | 0.157 | -0.004 | 7.18E-04 |
| 4 | 2.13E-03 | 9.07E-05 | 1.031 | 1.98E-03 | 9.90E-05 | 1.022 | 2.09E-03 | 9.18E-05 | 1.024 | 9.64E-04 | 0.000 | 0.000 | 1.16E-03 | 0.003 | -0.009 | 8.67E-04 | 0.055 | 0.002 | 8.67E-04 |
| 5 | 2.27E-03 | 8.50E-05 | 1.032 | 1.94E-03 | 1.02E-04 | 1.023 | 1.93E-03 | 9.63E-05 | 1.034 | 1.14E-03 | -0.004 | 0.005 | 1.11E-03 | 0.003 | -0.008 | 8.29E-04 | 0.106 | 0.000 | 8.29E-04 |
| 6 | 2.13E-03 | 8.84E-05 | 1.037 | 1.87E-03 | 1.05E-04 | 1.021 | 1.91E-03 | 9.72E-05 | 1.031 | 1.09E-03 | 0.000 | 0.000 | 1.00E-03 | 0.002 | -0.005 | 8.04E-04 | 0.096 | 0.001 | 6.45E-04 |
| 7 | 2.10E-03 | 9.00E-05 | 1.035 | 1.86E-03 | 1.04E-04 | 1.024 | 1.99E-03 | 9.25E-05 | 1.030 | 1.01E-03 | 0.000 | 0.000 | 1.05E-03 | 0.002 | -0.007 | 7.72E-04 | 0.049 | 0.004 | 7.72E-04 |
| 8 | 2.21E-03 | 8.80E-05 | 1.028 | 1.88E-03 | 1.02E-04 | 1.029 | 1.95E-03 | 9.53E-05 | 1.031 | 1.10E-03 | 0.001 | 0.004 | 1.01E-03 | 0.001 | -0.001 | 5.85E-04 | 0.208 | 0.000 | 5.85E-04 |
| 9 | 2.19E-03 | 8.81E-05 | 1.028 | 1.85E-03 | 1.00E-04 | 1.032 | 1.98E-03 | 9.27E-05 | 1.033 | 1.22E-03 | 0.000 | 0.000 | 9.99E-04 | 0.002 | -0.006 | 8.49E-04 | 0.059 | 0.002 | 8.49E-04 |
| 10 | 1.96E-03 | 1.00E-04 | 1.030 | 1.85E-03 | 1.05E-04 | 1.025 | 1.90E-03 | 9.79E-05 | 1.029 | 9.67E-04 | 0.000 | 0.000 | 9.25E-04 | 0.003 | -0.008 | 7.45E-04 | 0.097 | 0.000 | 7.45E-04 |
| 11 | 2.21E-03 | 8.58E-05 | 1.034 | 1.88E-03 | 1.02E-04 | 1.028 | 2.00E-03 | 9.49E-05 | 1.025 | 1.18E-03 | -0.004 | 0.005 | 1.11E-03 | 0.003 | -0.010 | 8.22E-04 | 0.091 | 0.001 | 8.22E-04 |
| 12 | 2.02E-03 | 9.83E-05 | 1.029 | 1.90E-03 | 1.04E-04 | 1.019 | 2.07E-03 | 9.30E-05 | 1.024 | 1.04E-03 | 0.000 | 0.000 | 1.11E-03 | 0.004 | -0.013 | 8.08E-04 | 0.089 | 0.001 | 8.08E-04 |
| 13 | 2.11E-03 | 9.28E-05 | 1.030 | 1.88E-03 | 1.02E-04 | 1.025 | 2.05E-03 | 9.57E-05 | 1.023 | 9.90E-04 | 0.000 | 0.000 | 1.06E-03 | 0.002 | -0.008 | 8.82E-04 | 0.047 | 0.003 | 8.82E-04 |
| 14 | 2.24E-03 | 8.86E-05 | 1.024 | 1.88E-03 | 1.05E-04 | 1.025 | 1.85E-03 | 1.01E-04 | 1.030 | 1.15E-03 | 0.000 | 0.000 | 1.01E-03 | 0.001 | -0.006 | 7.46E-04 | 0.100 | 0.001 | 5.64E-04 |
| 15* | 2.14E-03 | 8.82E-05 | 1.036 | 1.85E-03 | 1.04E-04 | 1.029 | 1.95E-03 | 9.86E-05 | 1.027 | 1.01E-03 | 0.000 | 0.000 | 1.06E-03 | 0.005 | -0.012 | 8.08E-04 | 0.084 | 0.002 | 8.08E-04 |
| 16 | 2.35E-03 | 8.55E-05 | 1.020 | 1.89E-03 | 1.00E-04 | 1.030 | 1.96E-03 | 9.39E-05 | 1.035 | 1.14E-03 | 0.000 | 0.001 | 1.14E-03 | 0.006 | -0.008 | 8.24E-04 | 0.106 | 0.001 | 8.24E-04 |

**Table S2.** Parameter estimates for 16 independent MiXeR runs for the "ring" simulated scenario.
The first column show the index of the optimization run (1-16), with the run having the smallest deviation from the median overlap pattern (as described in the methods section) marked with asterisk. Other columns show univariate polygenicities ($\pi_1^u, \pi_2^u, \pi_3^u$), discoverabilities ($\sigma_1, \sigma_2, \sigma_3$) and residual variances ($\sigma_{01}, \sigma_{02}, \sigma_{03}$) for Trait 1, Trait 2 and Trait 3 respectively; pairwise (bivariate) genetic overlaps ($\pi_{12}^b, \pi_{13}^b, \pi_{23}^b$), correlations of effect sizes within each of the three pairwise overlaps ($\rho_{12}, \rho_{13}, \rho_{23}$) and correlations between residuals ($\rho_{012}, \rho_{013}, \rho_{023}$) for Trait 1 and Trait 2, Trait 1 and Trait 3 and Trait 2 and Trait 3 pairs respectively; the genetic overlap between all three phenotypes ($\pi_{123}$).

| Index | $\pi_1^u$ | $\sigma_2$ | $\sigma_{01}$ | $\pi_2^u$ | $\sigma_2$ | $\sigma_{02}$ | $\pi_3^u$ | $\sigma_3$ | $\sigma_{03}$ | $\pi_{12}^b$ | $\rho_{12}$ | $\rho_{012}$ | $\pi_{13}^b$ | $\rho_{13}$ | $\rho_{013}$ | $\pi_{23}^b$ | $\rho_{23}$ | $\rho_{023}$ | $\pi_{123}$ |
|---|---|---|---|---|---|---|---|---|---|---|---|---|---|---|---|---|---|---|---|
| 1 | 2.25E-03 | 8.67E-05 | 1.026 | 2.02E-03 | 9.59E-05 | 1.023 | 1.92E-03 | 9.66E-05 | 1.033 | 1.25E-03 | 0.000 | 0.000 | 9.00E-04 | 0.002 | -0.005 | 7.97E-04 | -0.001 | 0.009 | 2.57E-05 |
| 2 | 2.47E-03 | 8.17E-05 | 1.017 | 2.01E-03 | 9.97E-05 | 1.016 | 2.03E-03 | 9.53E-05 | 1.025 | 1.23E-03 | -0.001 | 0.005 | 1.03E-03 | 0.000 | 0.000 | 1.16E-03 | 0.000 | 0.000 | 3.74E-04 |
| 3 | 2.36E-03 | 8.46E-05 | 1.020 | 1.78E-03 | 1.12E-04 | 1.018 | 1.94E-03 | 9.96E-05 | 1.027 | 1.13E-03 | -0.005 | 0.006 | 1.01E-03 | 0.001 | -0.004 | 1.05E-03 | -0.003 | 0.004 | 3.93E-04 |
| 4 | 2.26E-03 | 8.64E-05 | 1.024 | 1.94E-03 | 1.04E-04 | 1.015 | 2.00E-03 | 9.60E-05 | 1.026 | 1.21E-03 | 0.000 | 0.000 | 1.00E-03 | 0.001 | -0.002 | 1.04E-03 | -0.004 | 0.006 | 3.06E-04 |
| 5* | 2.22E-03 | 8.57E-05 | 1.029 | 1.88E-03 | 1.09E-04 | 1.012 | 1.92E-03 | 9.95E-05 | 1.027 | 1.10E-03 | -0.001 | 0.007 | 9.59E-04 | 0.000 | 0.000 | 1.08E-03 | 0.000 | 0.005 | 3.00E-04 |
| 6 | 2.32E-03 | 8.55E-05 | 1.023 | 1.88E-03 | 1.04E-04 | 1.024 | 1.90E-03 | 9.80E-05 | 1.034 | 1.23E-03 | -0.004 | 0.005 | 1.04E-03 | 0.001 | -0.005 | 1.14E-03 | -0.002 | 0.006 | 4.89E-04 |
| 7 | 2.20E-03 | 8.61E-05 | 1.032 | 1.95E-03 | 1.05E-04 | 1.012 | 2.07E-03 | 9.40E-05 | 1.021 | 1.26E-03 | -0.004 | 0.006 | 9.28E-04 | 0.003 | -0.004 | 1.11E-03 | -0.004 | 0.005 | 4.21E-04 |
| 8 | 2.33E-03 | 8.28E-05 | 1.026 | 1.94E-03 | 1.04E-04 | 1.018 | 2.05E-03 | 9.23E-05 | 1.031 | 1.12E-03 | 0.000 | 0.000 | 1.08E-03 | 0.000 | 0.000 | 1.07E-03 | -0.008 | 0.008 | 2.52E-04 |
| 9 | 2.24E-03 | 8.76E-05 | 1.024 | 1.85E-03 | 1.06E-04 | 1.021 | 1.97E-03 | 9.71E-05 | 1.027 | 1.12E-03 | -0.002 | 0.003 | 8.10E-04 | 0.001 | -0.005 | 1.01E-03 | -0.002 | 0.004 | 2.78E-04 |
| 10 | 2.23E-03 | 8.70E-05 | 1.027 | 1.84E-03 | 1.10E-04 | 1.016 | 2.11E-03 | 9.38E-05 | 1.021 | 1.12E-03 | -0.003 | 0.006 | 9.72E-04 | 0.002 | -0.004 | 1.09E-03 | -0.001 | 0.003 | 3.62E-04 |
| 11 | 2.27E-03 | 8.57E-05 | 1.026 | 1.87E-03 | 1.07E-04 | 1.019 | 1.89E-03 | 1.01E-04 | 1.028 | 1.17E-03 | -0.004 | 0.007 | 8.28E-04 | 0.002 | -0.005 | 9.69E-04 | 0.000 | 0.001 | 2.74E-04 |
| 12 | 2.15E-03 | 8.71E-05 | 1.034 | 1.93E-03 | 1.02E-04 | 1.019 | 2.05E-03 | 9.41E-05 | 1.025 | 1.24E-03 | -0.001 | 0.005 | 9.36E-04 | 0.000 | 0.000 | 1.11E-03 | -0.004 | 0.006 | 4.25E-04 |
| 13 | 2.26E-03 | 8.65E-05 | 1.023 | 1.82E-03 | 1.07E-04 | 1.024 | 1.93E-03 | 9.78E-05 | 1.029 | 1.16E-03 | -0.005 | 0.008 | 9.08E-04 | 0.002 | -0.005 | 9.69E-04 | -0.002 | 0.004 | 3.09E-04 |
| 14 | 2.28E-03 | 8.92E-05 | 1.021 | 1.99E-03 | 1.02E-04 | 1.013 | 2.00E-03 | 9.81E-05 | 1.024 | 1.11E-03 | -0.003 | 0.003 | 9.36E-04 | 0.002 | -0.005 | 1.05E-03 | 0.000 | 0.000 | 1.63E-04 |
| 15 | 2.15E-03 | 8.96E-05 | 1.028 | 1.99E-03 | 9.90E-05 | 1.016 | 1.92E-03 | 1.01E-04 | 1.026 | 1.16E-03 | -0.006 | 0.007 | 9.21E-04 | 0.000 | 0.000 | 1.05E-03 | -0.003 | 0.006 | 2.22E-04 |
| 16 | 2.19E-03 | 8.64E-05 | 1.032 | 1.88E-03 | 1.06E-04 | 1.017 | 1.96E-03 | 9.64E-05 | 1.031 | 1.14E-03 | -0.002 | 0.006 | 9.85E-04 | 0.002 | -0.005 | 1.09E-03 | -0.002 | 0.008 | 3.49E-04 |

**Table S3.** Parameter estimates for 16 independent MiXeR runs for the "equilibrium" simulated scenario.
The first column show the index of the optimization run (1-16), with the run having the smallest deviation from the median overlap pattern (as described in the methods section) marked with asterisk. Other columns show univariate polygenicities ($\pi_1^u, \pi_2^u, \pi_3^u$), discoverabilities ($\sigma_1, \sigma_2, \sigma_3$) and residual variances ($\sigma_{01}, \sigma_{02}, \sigma_{03}$) for Trait 1, Trait 2 and Trait 3 respectively; pairwise (bivariate) genetic overlaps ($\pi_{12}^b, \pi_{13}^b, \pi_{23}^b$), correlations of effect sizes within each of the three pairwise overlaps ($\rho_{12}, \rho_{13}, \rho_{23}$) and correlations between residuals ($\rho_{012}, \rho_{013}, \rho_{023}$) for Trait 1 and Trait 2, Trait 1 and Trait 3 and Trait 2 and Trait 3 pairs respectively; the genetic overlap between all three phenotypes ($\pi_{123}$).

| Index | $\pi_1^u$ | $\sigma_2$ | $\sigma_{01}$ | $\pi_2^u$ | $\sigma_2$ | $\sigma_{02}$ | $\pi_3^u$ | $\sigma_3$ | $\sigma_{03}$ | $\pi_{12}^b$ | $\rho_{12}$ | $\rho_{012}$ | $\pi_{13}^b$ | $\rho_{13}$ | $\rho_{013}$ | $\pi_{23}^b$ | $\rho_{23}$ | $\rho_{023}$ | $\pi_{123}$ |
|---|---|---|---|---|---|---|---|---|---|---|---|---|---|---|---|---|---|---|---|
| 1 | 1.99E-03 | 9.14E-05 | 1.035 | 1.95E-03 | 9.73E-05 | 1.027 | 2.02E-03 | 9.22E-05 | 1.030 | 1.01E-03 | 0.002 | -0.003 | 1.26E-03 | 0.002 | -0.003 | 9.45E-04 | -0.010 | 0.009 | 5.29E-04 |
| 2 | 2.11E-03 | 8.61E-05 | 1.032 | 2.02E-03 | 9.24E-05 | 1.030 | 1.97E-03 | 9.66E-05 | 1.026 | 1.04E-03 | 0.002 | -0.006 | 1.19E-03 | 0.002 | -0.006 | 8.77E-04 | -0.002 | 0.005 | 5.22E-04 |
| 3 | 2.04E-03 | 8.97E-05 | 1.034 | 2.12E-03 | 8.84E-05 | 1.027 | 2.07E-03 | 9.29E-05 | 1.026 | 1.15E-03 | 0.002 | -0.007 | 1.17E-03 | 0.000 | 0.000 | 1.06E-03 | -0.005 | 0.005 | 6.30E-04 |
| 4 | 2.10E-03 | 8.69E-05 | 1.035 | 2.06E-03 | 9.29E-05 | 1.027 | 1.97E-03 | 9.26E-05 | 1.034 | 1.01E-03 | 0.002 | -0.007 | 1.30E-03 | 0.002 | -0.004 | 9.15E-04 | -0.003 | 0.004 | 8.37E-04 |
| 5 | 2.13E-03 | 8.52E-05 | 1.032 | 2.03E-03 | 8.94E-05 | 1.036 | 1.88E-03 | 9.98E-05 | 1.029 | 1.24E-03 | 0.002 | -0.007 | 1.32E-03 | 0.001 | -0.001 | 7.70E-04 | -0.004 | 0.006 | 5.44E-04 |
| 6 | 2.09E-03 | 8.89E-05 | 1.030 | 1.98E-03 | 8.99E-05 | 1.038 | 2.12E-03 | 9.16E-05 | 1.022 | 1.05E-03 | 0.002 | -0.004 | 1.31E-03 | 0.000 | 0.000 | 8.53E-04 | 0.000 | 0.000 | 3.72E-04 |
| 7 | 2.16E-03 | 8.35E-05 | 1.034 | 1.96E-03 | 9.13E-05 | 1.038 | 2.17E-03 | 8.65E-05 | 1.031 | 1.09E-03 | 0.001 | -0.001 | 1.40E-03 | 0.001 | -0.003 | 9.22E-04 | 0.000 | 0.000 | 8.60E-04 |
| 8 | 2.06E-03 | 8.57E-05 | 1.039 | 2.00E-03 | 8.72E-05 | 1.042 | 1.92E-03 | 9.77E-05 | 1.033 | 1.18E-03 | 0.001 | -0.007 | 1.18E-03 | 0.001 | -0.001 | 9.62E-04 | -0.005 | 0.005 | 7.60E-04 |
| 9 | 1.98E-03 | 9.36E-05 | 1.030 | 2.12E-03 | 8.99E-05 | 1.025 | 2.11E-03 | 9.21E-05 | 1.024 | 1.18E-03 | 0.003 | -0.008 | 1.37E-03 | 0.001 | -0.002 | 1.03E-03 | -0.005 | 0.005 | 7.87E-04 |
| 10 | 1.87E-03 | 9.55E-05 | 1.040 | 2.02E-03 | 9.03E-05 | 1.035 | 2.06E-03 | 9.15E-05 | 1.031 | 9.49E-04 | 0.002 | -0.007 | 1.15E-03 | 0.000 | -0.001 | 1.02E-03 | -0.010 | 0.010 | 4.94E-04 |
| 11* | 2.00E-03 | 8.86E-05 | 1.039 | 2.05E-03 | 9.12E-05 | 1.026 | 1.99E-03 | 9.46E-05 | 1.028 | 1.14E-03 | 0.002 | -0.005 | 1.23E-03 | 0.000 | 0.000 | 8.84E-04 | -0.003 | 0.006 | 5.56E-04 |
| 12 | 2.10E-03 | 8.70E-05 | 1.034 | 2.07E-03 | 9.00E-05 | 1.027 | 2.03E-03 | 9.31E-05 | 1.030 | 1.03E-03 | 0.003 | -0.008 | 1.30E-03 | 0.005 | -0.010 | 8.93E-04 | -0.003 | 0.006 | 5.21E-04 |
| 13 | 2.03E-03 | 8.82E-05 | 1.037 | 2.07E-03 | 9.01E-05 | 1.030 | 2.07E-03 | 9.15E-05 | 1.031 | 1.14E-03 | 0.001 | -0.006 | 1.37E-03 | 0.002 | -0.006 | 9.13E-04 | -0.002 | 0.004 | 7.33E-04 |
| 14 | 2.06E-03 | 8.56E-05 | 1.041 | 2.01E-03 | 9.00E-05 | 1.034 | 2.15E-03 | 9.10E-05 | 1.023 | 1.12E-03 | 0.006 | -0.008 | 1.31E-03 | 0.002 | -0.008 | 9.41E-04 | -0.003 | 0.003 | 5.74E-04 |
| 15 | 2.04E-03 | 8.67E-05 | 1.038 | 2.05E-03 | 8.85E-05 | 1.033 | 2.12E-03 | 9.33E-05 | 1.020 | 1.16E-03 | 0.002 | -0.007 | 1.24E-03 | 0.000 | -0.004 | 9.75E-04 | -0.010 | 0.010 | 7.01E-04 |
| 16 | 2.13E-03 | 8.48E-05 | 1.034 | 2.23E-03 | 8.62E-05 | 1.023 | 1.92E-03 | 9.88E-05 | 1.028 | 1.13E-03 | 0.005 | -0.013 | 1.24E-03 | 0.000 | 0.000 | 9.26E-04 | -0.004 | 0.003 | 5.97E-04 |

**Table S4.** Parameter estimates for 16 independent MiXeR runs for type 2 diabetes (T2D), high-density lipoprotein (HDL) and estimated glomerular filtration rate (eGFR). The first column show the index of the optimization run (1-16), with the run having the smallest deviation from the median overlap pattern (as described in the methods section) marked with asterisk. Other columns show univariate polygenicities ($\pi_1^u, \pi_2^u, \pi_3^u$), discoverabilities ($\sigma_1, \sigma_2, \sigma_3$) and residual variances ($\sigma_{01}, \sigma_{02}, \sigma_{03}$) for T2D, HDL and eGFR respectively; pairwise (bivariate) genetic overlaps ($\pi_{12}^b, \pi_{13}^b, \pi_{23}^b$), correlations of effect sizes within each of the three pairwise overlaps ($\rho_{12}, \rho_{13}, \rho_{23}$) and correlations between residuals ($\rho_{012}, \rho_{013}, \rho_{023}$) for T2D and HDL, T2D and eGFR and HDL and eGFR pairs respectively; the genetic overlap between all three phenotypes ($\pi_{123}$).

| Index | $\pi_1^u$ | $\sigma_2$ | $\sigma_{01}$ | $\pi_2^u$ | $\sigma_2$ | $\sigma_{02}$ | $\pi_3^u$ | $\sigma_3$ | $\sigma_{03}$ | $\pi_{12}^b$ | $\rho_{12}$ | $\rho_{012}$ | $\pi_{13}^b$ | $\rho_{13}$ | $\rho_{013}$ | $\pi_{23}^b$ | $\rho_{23}$ | $\rho_{023}$ | $\pi_{123}$ |
|---|---|---|---|---|---|---|---|---|---|---|---|---|---|---|---|---|---|---|---|
| 1 | 5.18E-04 | 1.30E-04 | 1.066 | 3.62E-04 | 1.80E-04 | 1.276 | 4.16E-04 | 8.45E-05 | 1.058 | 2.75E-04 | -0.411 | -0.228 | 1.52E-04 | 0.217 | 0.000 | 1.51E-04 | 0.096 | 0.003 | 1.35E-04 |
| 2* | 5.16E-04 | 1.30E-04 | 1.066 | 3.81E-04 | 1.69E-04 | 1.268 | 4.27E-04 | 8.31E-05 | 1.058 | 2.66E-04 | -0.429 | -0.227 | 1.70E-04 | 0.221 | 0.000 | 1.60E-04 | 0.146 | 0.003 | 1.24E-04 |
| 3 | 4.58E-04 | 1.48E-04 | 1.067 | 3.71E-04 | 1.75E-04 | 1.276 | 4.03E-04 | 8.50E-05 | 1.065 | 1.68E-04 | -0.679 | -0.219 | 1.64E-04 | 0.077 | 0.000 | 1.02E-04 | 0.218 | 0.000 | 1.02E-04 |
| 4 | 4.85E-04 | 1.39E-04 | 1.066 | 3.87E-04 | 1.68E-04 | 1.268 | 3.85E-04 | 9.01E-05 | 1.065 | 2.78E-04 | -0.503 | -0.222 | 1.52E-04 | 0.156 | 0.000 | 1.44E-04 | 0.128 | 0.001 | 1.31E-04 |
| 5 | 4.83E-04 | 1.39E-04 | 1.066 | 4.06E-04 | 1.57E-04 | 1.255 | 4.05E-04 | 8.74E-05 | 1.061 | 3.22E-04 | -0.225 | -0.219 | 1.39E-04 | 0.216 | 0.000 | 1.54E-04 | 0.154 | 0.000 | 1.05E-04 |
| 6 | 4.75E-04 | 1.37E-04 | 1.079 | 3.70E-04 | 1.74E-04 | 1.276 | 3.96E-04 | 8.81E-05 | 1.066 | 2.55E-04 | -0.300 | -0.210 | 2.09E-04 | 0.077 | 0.000 | 1.20E-04 | 0.150 | 0.000 | 1.20E-04 |
| 7 | 4.98E-04 | 1.34E-04 | 1.066 | 3.72E-04 | 1.73E-04 | 1.276 | 4.01E-04 | 8.36E-05 | 1.072 | 1.77E-04 | -0.764 | -0.222 | 1.59E-04 | 0.146 | 0.000 | 1.52E-04 | 0.144 | 0.000 | 5.57E-05 |
| 8 | 5.21E-04 | 1.31E-04 | 1.066 | 3.78E-04 | 1.65E-04 | 1.281 | 4.43E-04 | 7.93E-05 | 1.057 | 2.73E-04 | -0.394 | -0.223 | 1.79E-04 | 0.090 | 0.002 | 1.49E-04 | 0.220 | 0.000 | 1.27E-04 |
| 9 | 5.04E-04 | 1.34E-04 | 1.066 | 3.70E-04 | 1.75E-04 | 1.276 | 4.11E-04 | 8.39E-05 | 1.064 | 1.65E-04 | -0.764 | -0.217 | 1.65E-04 | 0.139 | 0.000 | 1.46E-04 | 0.078 | 0.000 | 7.53E-05 |
| 10 | 5.12E-04 | 1.31E-04 | 1.066 | 3.83E-04 | 1.66E-04 | 1.276 | 4.03E-04 | 8.75E-05 | 1.062 | 1.77E-04 | -0.896 | -0.222 | 1.53E-04 | 0.213 | 0.000 | 1.42E-04 | 0.219 | 0.000 | 7.32E-05 |
| 11 | 5.47E-04 | 1.30E-04 | 1.048 | 3.75E-04 | 1.70E-04 | 1.276 | 4.19E-04 | 8.44E-05 | 1.058 | 2.21E-04 | -0.618 | -0.222 | 1.28E-04 | 0.216 | 0.000 | 1.53E-04 | 0.155 | 0.001 | 1.28E-04 |
| 12 | 4.94E-04 | 1.30E-04 | 1.078 | 3.69E-04 | 1.70E-04 | 1.276 | 4.05E-04 | 8.71E-05 | 1.062 | 2.14E-04 | -0.529 | -0.209 | 1.49E-04 | 0.223 | 0.000 | 1.42E-04 | 0.156 | 0.000 | 3.80E-05 |
| 13 | 4.95E-04 | 1.37E-04 | 1.065 | 3.80E-04 | 1.67E-04 | 1.276 | 3.98E-04 | 8.58E-05 | 1.068 | 1.76E-04 | -0.777 | -0.222 | 1.55E-04 | 0.151 | 0.000 | 1.49E-04 | 0.074 | 0.001 | 1.00E-06 |
| 14 | 4.61E-04 | 1.44E-04 | 1.072 | 3.81E-04 | 1.67E-04 | 1.276 | 4.00E-04 | 8.51E-05 | 1.070 | 1.48E-04 | -0.847 | -0.223 | 1.41E-04 | 0.220 | 0.000 | 2.04E-04 | 0.151 | 0.000 | 1.01E-04 |
| 15 | 4.60E-04 | 1.48E-04 | 1.069 | 3.66E-04 | 1.79E-04 | 1.276 | 3.93E-04 | 8.83E-05 | 1.065 | 2.73E-04 | -0.322 | -0.219 | 1.53E-04 | 0.144 | 0.000 | 1.14E-04 | 0.150 | 0.000 | 8.10E-05 |
| 16 | 4.54E-04 | 1.52E-04 | 1.066 | 3.77E-04 | 1.70E-04 | 1.276 | 4.01E-04 | 8.84E-05 | 1.055 | 2.93E-04 | -0.439 | -0.218 | 1.53E-04 | -0.006 | 0.004 | 1.60E-04 | 0.065 | 0.006 | 8.68E-05 |

**Table S5.** Parameter estimates for 16 independent MiXeR runs for ulcerative colitis (UC), psoriasis (PS), and multiple sclerosis (MS).
The first column show the index of the optimization run (1-16), with the run having the smallest deviation from the median overlap pattern (as described in the methods section) marked with asterisk. Other columns show univariate polygenicities ($\pi_1^u, \pi_2^u, \pi_3^u$), discoverabilities ($\sigma_1, \sigma_2, \sigma_3$) and residual variances ($\sigma_{01}, \sigma_{02}, \sigma_{03}$) for UC, PS and MS respectively; pairwise (bivariate) genetic overlaps ($\pi_{12}^b, \pi_{13}^b, \pi_{23}^b$), correlations of effect sizes within each of the three pairwise overlaps ($\rho_{12}, \rho_{13}, \rho_{23}$) and correlations between residuals ($\rho_{012}, \rho_{013}, \rho_{023}$) for UC and PS, UC and MS and PS and MS pairs respectively; the genetic overlap between all three phenotypes ($\pi_{123}$).

| Index | $\pi_1^u$ | $\sigma_2$ | $\sigma_{01}$ | $\pi_2^u$ | $\sigma_2$ | $\sigma_{02}$ | $\pi_3^u$ | $\sigma_3$ | $\sigma_{03}$ | $\pi_{12}^b$ | $\rho_{12}$ | $\rho_{012}$ | $\pi_{13}^b$ | $\rho_{13}$ | $\rho_{013}$ | $\pi_{23}^b$ | $\rho_{23}$ | $\rho_{023}$ | $\pi_{123}$ |
|---|---|---|---|---|---|---|---|---|---|---|---|---|---|---|---|---|---|---|---|
| 1 | 1.22E-04 | 1.23E-03 | 1.121 | 1.06E-04 | 7.62E-04 | 1.095 | 2.02E-04 | 8.39E-04 | 1.043 | 6.32E-05 | 0.192 | 0.011 | 8.12E-05 | 0.185 | 0.068 | 8.15E-05 | 0.222 | 0.000 | 6.32E-05 |
| 2 | 1.37E-04 | 1.12E-03 | 1.118 | 1.04E-04 | 7.75E-04 | 1.093 | 1.98E-04 | 8.83E-04 | 1.043 | 6.70E-05 | 0.156 | 0.011 | 9.18E-05 | 0.217 | 0.066 | 7.96E-05 | 0.212 | 0.000 | 6.70E-05 |
| 3 | 1.51E-04 | 1.01E-03 | 1.117 | 1.07E-04 | 7.55E-04 | 1.094 | 1.82E-04 | 9.07E-04 | 1.049 | 6.80E-05 | 0.227 | 0.010 | 9.09E-05 | 0.212 | 0.064 | 8.33E-05 | 0.184 | 0.004 | 6.80E-05 |
| 4 | 1.17E-04 | 1.27E-03 | 1.122 | 1.40E-04 | 6.32E-04 | 1.087 | 1.87E-04 | 9.00E-04 | 1.047 | 6.59E-05 | 0.200 | 0.015 | 7.57E-05 | 0.224 | 0.064 | 9.31E-05 | 0.211 | 0.000 | 6.59E-05 |
| 5 | 1.15E-04 | 1.31E-03 | 1.121 | 8.61E-05 | 8.94E-04 | 1.095 | 1.90E-04 | 9.28E-04 | 1.043 | 5.68E-05 | 0.140 | 0.012 | 7.92E-05 | 0.203 | 0.064 | 6.82E-05 | 0.220 | 0.000 | 5.68E-05 |
| 6 | 1.33E-04 | 1.15E-03 | 1.117 | 8.46E-05 | 8.81E-04 | 1.096 | 1.96E-04 | 8.61E-04 | 1.047 | 5.76E-05 | 0.138 | 0.014 | 8.98E-05 | 0.207 | 0.065 | 6.92E-05 | 0.180 | 0.004 | 5.76E-05 |
| 7 | 1.26E-04 | 1.21E-03 | 1.116 | 1.45E-04 | 5.89E-04 | 1.089 | 2.06E-04 | 8.40E-04 | 1.046 | 7.57E-05 | 0.215 | 0.009 | 8.19E-05 | 0.238 | 0.063 | 9.90E-05 | 0.231 | 0.000 | 7.57E-05 |
| 8 | 1.06E-04 | 1.35E-03 | 1.126 | 1.15E-04 | 7.15E-04 | 1.089 | 1.87E-04 | 9.06E-04 | 1.045 | 6.37E-05 | 0.205 | 0.011 | 6.89E-05 | 0.209 | 0.066 | 8.40E-05 | 0.203 | 0.001 | 6.37E-05 |
| 9 | 1.14E-04 | 1.25E-03 | 1.124 | 1.06E-04 | 7.40E-04 | 1.094 | 2.17E-04 | 8.16E-04 | 1.039 | 6.87E-05 | 0.174 | 0.005 | 8.06E-05 | 0.205 | 0.065 | 8.43E-05 | 0.165 | 0.001 | 6.87E-05 |
| 10* | 1.20E-04 | 1.23E-03 | 1.122 | 1.17E-04 | 6.88E-04 | 1.090 | 2.02E-04 | 8.71E-04 | 1.038 | 6.18E-05 | 0.199 | 0.010 | 7.81E-05 | 0.249 | 0.064 | 9.06E-05 | 0.169 | 0.001 | 6.18E-05 |
| 11 | 1.20E-04 | 1.23E-03 | 1.119 | 1.22E-04 | 6.54E-04 | 1.093 | 1.87E-04 | 8.68E-04 | 1.051 | 7.43E-05 | 0.000 | 0.014 | 8.31E-05 | 0.221 | 0.065 | 9.43E-05 | 0.161 | 0.002 | 7.43E-05 |
| 12 | 1.18E-04 | 1.28E-03 | 1.121 | 1.36E-04 | 6.30E-04 | 1.091 | 1.99E-04 | 8.53E-04 | 1.045 | 8.06E-05 | 0.197 | 0.001 | 7.73E-05 | 0.225 | 0.067 | 8.97E-05 | 0.209 | 0.000 | 7.73E-05 |
| 13 | 1.11E-04 | 1.33E-03 | 1.125 | 1.03E-04 | 7.87E-04 | 1.094 | 1.69E-04 | 1.03E-03 | 1.048 | 6.11E-05 | 0.152 | 0.015 | 7.06E-05 | 0.234 | 0.064 | 3.17E-05 | 0.439 | 0.000 | 3.17E-05 |
| 14 | 1.21E-04 | 1.22E-03 | 1.120 | 1.09E-04 | 6.98E-04 | 1.095 | 1.69E-04 | 1.01E-03 | 1.049 | 7.10E-05 | 0.188 | 0.015 | 6.36E-05 | 0.354 | 0.067 | 8.73E-05 | 0.179 | 0.001 | 6.15E-05 |
| 15 | 1.15E-04 | 1.28E-03 | 1.125 | 1.21E-04 | 6.71E-04 | 1.092 | 2.08E-04 | 8.60E-04 | 1.042 | 7.26E-05 | 0.230 | 0.008 | 8.56E-05 | 0.214 | 0.066 | 9.93E-05 | 0.175 | 0.003 | 7.26E-05 |
| 16 | 1.19E-04 | 1.23E-03 | 1.123 | 1.12E-04 | 7.14E-04 | 1.091 | 2.16E-04 | 8.01E-04 | 1.044 | 6.94E-05 | 0.152 | 0.012 | 9.23E-05 | 0.212 | 0.061 | 9.17E-05 | 0.211 | 0.001 | 6.94E-05 |

**Table S6.** Parameter estimates for 16 independent MiXeR runs for placental weight (PW), schizophrenia (SCZ) and height.
The first column show the index of the optimization run (1-16), with the run having the smallest deviation from the median overlap pattern (as described in the methods section) marked with asterisk. Other columns show univariate polygenicities ($\pi_1^u, \pi_2^u, \pi_3^u$), discoverabilities ($\sigma_1, \sigma_2, \sigma_3$) and residual variances ($\sigma_{01}, \sigma_{02}, \sigma_{03}$) for PW, SCZ and height respectively; pairwise (bivariate) genetic overlaps ($\pi_{12}^b, \pi_{13}^b, \pi_{23}^b$), correlations of effect sizes within each of the three pairwise overlaps ($\rho_{12}, \rho_{13}, \rho_{23}$) and correlations between residuals ($\rho_{012}, \rho_{013}, \rho_{023}$) for PW and SCZ, PW and height and SCZ and height pairs respectively; the genetic overlap between all three phenotypes ($\pi_{123}$).

| Index | $\pi_1^u$ | $\sigma_2$ | $\sigma_{01}$ | $\pi_2^u$ | $\sigma_2$ | $\sigma_{02}$ | $\pi_3^u$ | $\sigma_3$ | $\sigma_{03}$ | $\pi_{12}^b$ | $\rho_{12}$ | $\rho_{012}$ | $\pi_{13}^b$ | $\rho_{13}$ | $\rho_{013}$ | $\pi_{23}^b$ | $\rho_{23}$ | $\rho_{023}$ | $\pi_{123}$ |
|---|---|---|---|---|---|---|---|---|---|---|---|---|---|---|---|---|---|---|---|
| 1* | 3.91E-04 | 1.74E-04 | 1.038 | 2.88E-03 | 6.06E-05 | 1.155 | 1.28E-03 | 1.81E-04 | 2.096 | 7.11E-05 | -0.233 | 0.000 | 2.83E-04 | 0.557 | 0.037 | 2.80E-04 | -0.157 | -0.023 | 7.11E-05 |
| 2 | 3.93E-04 | 1.76E-04 | 1.035 | 2.95E-03 | 5.94E-05 | 1.158 | 1.26E-03 | 1.82E-04 | 2.126 | 1.08E-04 | 0.002 | -0.005 | 2.95E-04 | 0.531 | 0.038 | 2.42E-04 | -0.311 | -0.011 | 1.08E-04 |
| 3 | 3.89E-04 | 1.72E-04 | 1.042 | 2.81E-03 | 6.21E-05 | 1.157 | 1.27E-03 | 1.83E-04 | 2.108 | 4.81E-05 | -0.228 | -0.008 | 3.10E-04 | 0.510 | 0.035 | 3.04E-04 | -0.145 | -0.017 | 3.72E-05 |
| 4 | 4.19E-04 | 1.62E-04 | 1.040 | 2.93E-03 | 6.13E-05 | 1.145 | 1.25E-03 | 1.86E-04 | 2.122 | 1.51E-05 | 1.000 | -0.006 | 3.09E-04 | 0.507 | 0.039 | 3.04E-04 | -0.151 | -0.016 | 1.00E-06 |
| 5 | 3.98E-04 | 1.71E-04 | 1.037 | 2.85E-03 | 6.19E-05 | 1.153 | 1.26E-03 | 1.86E-04 | 2.106 | 8.03E-05 | 0.002 | -0.008 | 2.96E-04 | 0.540 | 0.037 | 2.98E-04 | -0.166 | -0.012 | 8.03E-05 |
| 6 | 3.68E-04 | 1.77E-04 | 1.045 | 2.84E-03 | 6.15E-05 | 1.158 | 1.24E-03 | 1.86E-04 | 2.125 | 7.57E-05 | 0.004 | -0.004 | 2.70E-04 | 0.556 | 0.038 | 3.04E-04 | -0.135 | -0.020 | 7.57E-05 |
| 7 | 4.15E-04 | 1.63E-04 | 1.041 | 2.81E-03 | 6.26E-05 | 1.154 | 1.26E-03 | 1.84E-04 | 2.105 | 8.05E-06 | 0.889 | -0.001 | 3.01E-04 | 0.549 | 0.035 | 2.57E-04 | -0.296 | -0.002 | 1.00E-06 |
| 8 | 4.13E-04 | 1.66E-04 | 1.039 | 2.87E-03 | 6.16E-05 | 1.153 | 1.27E-03 | 1.82E-04 | 2.099 | 7.34E-05 | -0.229 | 0.000 | 3.03E-04 | 0.516 | 0.038 | 2.79E-04 | -0.289 | -0.002 | 7.34E-05 |
| 9 | 3.56E-04 | 1.80E-04 | 1.044 | 2.88E-03 | 6.09E-05 | 1.154 | 1.23E-03 | 1.89E-04 | 2.137 | 9.62E-05 | 0.003 | -0.006 | 2.70E-04 | 0.545 | 0.038 | 3.13E-04 | -0.224 | 0.000 | 9.62E-05 |
| 10 | 4.16E-04 | 1.61E-04 | 1.039 | 2.95E-03 | 5.97E-05 | 1.151 | 1.26E-03 | 1.84E-04 | 2.111 | 5.31E-05 | -0.267 | -0.004 | 3.06E-04 | 0.535 | 0.036 | 2.44E-04 | -0.349 | -0.004 | 5.31E-05 |
| 11 | 3.69E-04 | 1.85E-04 | 1.038 | 2.98E-03 | 5.91E-05 | 1.154 | 1.25E-03 | 1.86E-04 | 2.126 | 2.25E-05 | 0.775 | -0.008 | 2.77E-04 | 0.508 | 0.038 | 3.17E-04 | -0.123 | -0.016 | 1.21E-05 |
| 12 | 3.63E-04 | 1.84E-04 | 1.041 | 2.78E-03 | 6.25E-05 | 1.158 | 1.23E-03 | 1.88E-04 | 2.125 | 9.21E-05 | 0.003 | -0.006 | 2.74E-04 | 0.538 | 0.038 | 2.89E-04 | -0.144 | -0.020 | 9.21E-05 |
| 13 | 3.94E-04 | 1.69E-04 | 1.041 | 2.78E-03 | 6.27E-05 | 1.159 | 1.26E-03 | 1.83E-04 | 2.117 | 1.02E-04 | 0.003 | -0.008 | 2.99E-04 | 0.527 | 0.033 | 2.10E-04 | -0.357 | -0.013 | 1.02E-04 |
| 14 | 3.30E-04 | 1.93E-04 | 1.045 | 3.10E-03 | 5.74E-05 | 1.148 | 1.24E-03 | 1.87E-04 | 2.125 | 1.57E-05 | 0.969 | -0.006 | 2.65E-04 | 0.542 | 0.037 | 3.51E-04 | -0.145 | -0.009 | 1.00E-06 |
| 15 | 3.89E-04 | 1.73E-04 | 1.040 | 2.86E-03 | 6.08E-05 | 1.156 | 1.26E-03 | 1.84E-04 | 2.090 | 9.51E-05 | 0.002 | -0.006 | 2.91E-04 | 0.534 | 0.036 | 1.08E-04 | -0.652 | -0.016 | 9.51E-05 |
| 16 | 3.68E-04 | 1.78E-04 | 1.043 | 2.90E-03 | 5.98E-05 | 1.159 | 1.26E-03 | 1.82E-04 | 2.117 | 1.41E-05 | 0.882 | -0.004 | 2.78E-04 | 0.507 | 0.039 | 2.82E-04 | -0.167 | -0.014 | 1.70E-06 |
